# Opioid Overdose and Naloxone Administration Knowledge and Perceived Competency in A Probability Sample of Indiana Urban Communities with Large Black Populations

**DOI:** 10.1101/2024.11.06.24316851

**Authors:** Shin Hyung Lee, Jon Agley, Vatsla Sharma, Francesca Williamson, Pengyue Zhang, Dong-Chul Seo

## Abstract

**Background:** Despite widespread naloxone distribution efforts, opioid-involved overdose rates remain high, with rates in recent years indicating disproportionate increases in the death rate for Black residents. This study evaluated knowledge and perceived competency regarding opioid overdose and naloxone administration among urban Indiana residents.

**Methods:** As part of a federally funded project (#CPIMP221346), the Indiana University Center for Survey Research conducted community probability surveys from March to May 2023, sampling 772 residents in Indiana’s urban communities across 8 zip code areas. The study explored individual and community level factors affecting residents’ knowledge and perceived competency regarding opioid overdose and naloxone administration.

**Results:** Multilevel analysis identified race, sex, household income, education, length of time lived in community, and a history of opioid overdose as significant predictors of knowledge. Participants who identified as White scored a mean of 6.65 out of 10, those of Other races scored 5.75, and Black participants scored 5.70, on a 10 item questionnaire developed from the Overdose Knowledge Scale (OOKS) and the Opioid Overdose Attitudes Scale (OOAS) (*p* < 0.001). Additionally, there was significant cross-level interaction between race and poverty, such that Black residents living in poorer neighborhoods displayed lower knowledge scores than White counterparts (□ = 1.06, *p* = 0.039). However, in terms of perceived competency, only age and a history of opioid overdose, were found to be significant predictors.

**Conclusions:** These findings underscore the importance of community education to increase knowledge and perceived competency regarding opioid overdose and naloxone administration among Black populations particularly living in socioeconomically disadvantated areas.

## Introduction

With the recent proliferation of illicit synthetic opioids, including fentanyl and its analogues, Indiana, like other states, is facing a significant public health challenge from opioid-involved overdoses (1, 2). In 2022, Indiana ranked as the 13^th^ highest state in drug overdose mortality nationwide, with a rate of 41.0 deaths per 100,000 population. Moreover, Indiana recorded 2,072 deaths attributed to opioid overdoses, comprising 77% of the overall drug overdose fatalities within the state in 2022 (3). In Indianapolis, the most populous city in Indiana, there were 195 fentanyl-related drug overdose deaths in 2018, and this number had increased to 799 deaths in 2021 (1).

Researchers have expressed concern that the increase in opioid-involved overdoses and fatalities is disproportionately affecting marginalized communities, with Black Americans in urban settings particularly impacted (4–6). In 2018 and 2019, the age-adjusted opioid overdose death rate per 100,000 people was higher for White people (18.8 and 19.2, respectively) than for Black people (14.1 and 17.3, respectively), but in 2020, data began showing a higher death rate for Black people than White people and diverging trend lines. By 2022, the opioid overdose death rate for Black people was 36.6 and the rate for White people was 27.6 (3). Gondré-Lewis et al. (2023) also reported that several major urban areas, including Baltimore, Chicago, Detroit, and Philadelphia, with large Black populations, had markedly higher opioid-related death rates compared to rural areas (5). Similarly, Black communities in Indiana experienced the largest increase in opioid-related overdose deaths in recent years (7). However, the underlying reasons for these emerging patterns remain unclear. Hence, studies on opioid-involved overdoses and fatalities among Black populations in large urban areas are warranted.

Administration of naloxone to reverse opioid overdoses is one key area where data suggest population-level differences that may be associated with opioid-related death rates. Naloxone, an opioid antagonist, is a safe and effective medication that can be easily administered to anyone undergoing an opioid overdose to reverse that overdose (8, 9). Research suggests that the incidence of fatal opioid overdoses could decrease if community members have access to naloxone and are educated about opioid-involved overdose and naloxone administration (10–12). Yet, in their Baltimore city based study, Dayton et al. (2020) found that White participants had significantly higher odds of receiving naloxone training and using naloxone compared to Black participants (13). In their study on drug overdose deaths, Takemoto et al. (2024) and Ray et al. (2019) found racial disparities in naloxone administration, with White decedents more likely to have had naloxone administered compared to Black decedents (14, 15). At the same time, most U.S. states have enacted legislation to expand its accessibility and utilization among laypersons to address public health emergency of opioid-involved fatalities (8, 11, 16). In addition, the U.S. Food and Drug Administration recently approved some formulations of naloxone for over-the-counter sale nationwide (17).

The State of Indiana has undertaken substantial efforts to expand access to naloxone in recent years, and more than 578,000 doses of naloxone have been distributed throughout Indiana as part of these efforts since 2020 (18). Still, there are multiple factors that may hinder timely naloxone administration at overdose scenes, such as lack of knowledge about opioid overdose and naloxone administration methods (19). Such barriers to timely naloxone administration to reverse opioid-involved overdoses are complex (19) and may be related to insufficient understanding of opioid overdose and naloxone administration (20, 21). Courtney et al. (2023) found that lack of knowledge about opioid overdose and naloxone administration among laypersons were among the major barriers for using naloxone at overdose scenes (20). To the best of our knowledge, no study has evaluated knowledge and competency regarding opioid overdose and naloxone administration among Indiana residents, particularly among urban Black populations. Considering the current trends in opioid-involved overdoses and fatalities in Indiana, it is important to assess knowledge and competency regarding opioid overdose and naloxone administration in urban Black communities to identify factors that could possibly reverse these trends in Indiana.

The objectives of this study were (1) to measure the level of knowledge and perceived competency regarding opioid overdose and naloxone administration in recent probability samples of Black urban communities in Indiana, (2) investigate racial and other sociodemographic correlates of knowledge and perceived competency, and (3) assess possible relationships, if any, between community-level factors (e.g., proportion of Black residents in the community, community-level income and employment rates) and the knowledge and perceived competency.

## 2. Methods

### 2.1. Ethics statement

This study was approved by the Indiana University Institutional Review Board (IUIRB), which determined that the research met the requirements for federal and University criteria for exemption (approved protocol ID: 16953). Informed consent was obtained for all participants. Waiver of documentation of consent was approved by the IUIRB.

### 2.2. Sample and administration

This study was part of the Multi-Sector and Multi-Level Community-Driven Approaches to Remove Structural Racism and Overdose Deaths in Black Indianapolis Communities (MACRO-B), a federally funded project that includes opioid overdose and naloxone administration community education programming to reduce opioid overdose deaths in Black Indianapolis communities (#CPIMP221346). As part of the project, data were collected from March 2023 to May 2023 via mail and web-based household probability community surveys with samples of adults aged 18 years or older. Data collection was managed by the Indiana University Center for Survey Research. Probability sampling was used to select respondents who were representative of 8 zip code areas of urban communities in Indiana with Black populations greater than 40%: inner-city Indianapolis (46202, 46205, 46208, and 46218), Gary (46408), Merrillville (46410), South Bend (46628), and Fort Wayne (46806).

The study followed the guidelines outlined by the American Association for Public Opinion Research (22). The survey administration process involved several sequential stages. On day 1, a preliminary letter and a $1 pre-survey incentive were sent out, instructing the household member with the most recent birthday to complete the online survey. This communication included instructions for accessing the survey and highlighted a $20 incentive for completion. On day 10, reminder letters were sent to non-respondents, emphasizing the importance of participation and reminding them of the incentive. On day 20, a second reminder letter that also included a self-addressed and stamped envelope was sent to non-respondents, accompanied by a paper version of the survey, to those who had not responded to the online survey. Subsequently, on day 35, a third reminder letter and paper survey with self-addressed and stamped envelopes were dispatched to encourage participation from non-respondents. Finally, on day 50, a final reminder letter was sent, urging participation either online or through the paper survey, with mailing scheduled for day 55. The response rate was 23% using AAPOR response rate formula #4 (22).

### 2.3. Measures

#### 2.3.1. Knowledge on opioid overdose and naloxone administration

To assess knowledge, we utilized 10 questions drawn from two validated tools: the Opioid Overdose Knowledge Scale (OOKS) and the Opioid Overdose Attitudes Scale (OOAS). Detailed information about these tools can be found in Williams et al. (23). The questionnaire included 6 true/false items and 4 multiple-choice items designed to test basic knowledge about identifying the risks, signs, and symptoms of opioid overdose, as well as understanding the proper use of naloxone (see Table 2 for all items). Each correct answer earned one point, resulting in a possible score range of 0 to 10 for knowledge on opioid overdose and naloxone administration. We calculated mean scores by race, ensuring that all scores were weighted accordingly.

#### 2.3.2. Perceived competency to manage opioid overdoses

To assess the perceived level of competency in managing opioid overdoses, we asked the survey respondents to rate their agreement with 7 items on a Likert-type scale ranging from 1 to 5 (see Table 2 for the specific items). These items were also derived from the Opioid Overdose Knowledge Scale (OOKS) and the Opioid Overdose Attitudes Scale (OOAS) (23). Each item was scored individually, and appropriate reverse coding was made so that a higher score always indicated a higher perceived level of competency. We then calculated the weighted mean scores of all 7 items by race.

#### 2.3.3. Individual-level and zip code-level predictors

One of the key variables in this study was race. In analyses, variable responses were dummy coded as White, Black, and Other. Due to the small sample sizes for specific groups (Asian = 22, American Indian or Alaska Native = 21, and Native Hawaiian or Pacific Islander = 6), we combined those individuals with the “Other” category to ensure sufficient numbers for meaningful analyses (24). Geographic area was determined by the eight zip code areas, which were then dummy coded by region as inner-city Indianapolis (46202, 46205, 46208, and 46218) and other urban areas such as Gary (46408), Merrillville (46410), South Bend (46628), and Fort Wayne (46806). Several demographic factors were included as covariates: age group (18-24, 25-34, 35-44, 45-54, 55-64, 65-74, and 75+), biological sex at birth (women, men), ethnicity (Non-Latino, Latino, educational level (some college or higher, high school/GED or less), and household income which had the following categories: less than $15,000; $15,000–$24,999; $25,000–$34,999; $35,000–$49,999; $50,000–$74,999; $75,000–$99,999; and $100,000 or more. For analysis purposes, income was collapsed into three broader categories (over $100,000, $35,000-$99,999, less than $35,000). Other covariates included duration of residence in the community (more than 3 years, 1 to 3 years, less than a year) and history with opioid overdose (yes/no). The variable for opioid overdose history was defined by whether individual had personally experienced an opioid overdose or had a family member or close friend who died from an opioid overdose. Additionally, zip code-level factors were included such as proportion of Black residents, number of individuals with income below the 200 percent of federal poverty level (FPL<200%), median household income, proportion of individuals with high school or higher educational attainment among those who were 25 years or older, and employment rate. These factors were obtained from the 2022 American Community Survey 5-year estimates (25).

### 2.4. Statistical analysis

Out of the 772 survey respondents, 47 cases with missing information were excluded from the analysis using listwise deletion, resulting in a final analytic sample of 725 respondents. We used listwise deletion to handle missing data, as it is a standard method when the proportion of missing data is relatively low. In this study, only 6% of the cases were missing data, which is considered a low rate (26). Listwise deletion ensures that the final analytic sample remains consistent across variables, which helps to simplify the analysis and interpretation of results (27).

We calculated both unweighted frequencies and weighted proportions for respondents who answered the questions on opioid overdose and naloxone administration correctly, along with their demographic variables. Additionally, we computed weighted mean scores for each item to assess perceived competency in managing opioid overdoses. We then compared these responses across different racial groups. We built multilevel models to identify factors associated with two outcomes: (a) knowledge and (b) perceived competency regarding opioid overdose and naloxone administration. Multilevel analysis allows simultaneous examination of individual-level and zip code-level factors as predictors of our outcome variables, as an individual’s knowledge and perceived competency regarding opioid overdose and naloxone administration are likely to be associated with structural factors associated with their place of residence. Following Lee et al. (2015), we constructed nested models sequentially and tested model fit. We started with a null model, then added individual-level socioeconomic factors, followed by overdose experience, and finally included zip code-level predictors and cross-level interaction terms (28). Model comparison and fitness were assessed using the Bayesian Information Criterion (BIC), Akaike Information Criterion (AIC), and log likelihood. All analyses were conducted using R version 4.3.2 (29).

## 3. Results

### 3.1. Demographic characteristics

Table 1 provides a breakdown of the demographic and socioeconomic characteristics of the survey respondents categorized by race, including White (n = 359), Black (n = 347), and Other racial groups (n = 60). There were significant differences observed for the following variables: ethnicity (*p* = 0.002), age group (*p* < 0.001), educational attainment (*p* = 0.001), household income (*p* < 0.001), and opioid overdose history (*p* = 0.003). The variables of region, sex, and length of time lived in the communities did not show statistically significant differences by race.

**Table 1.**
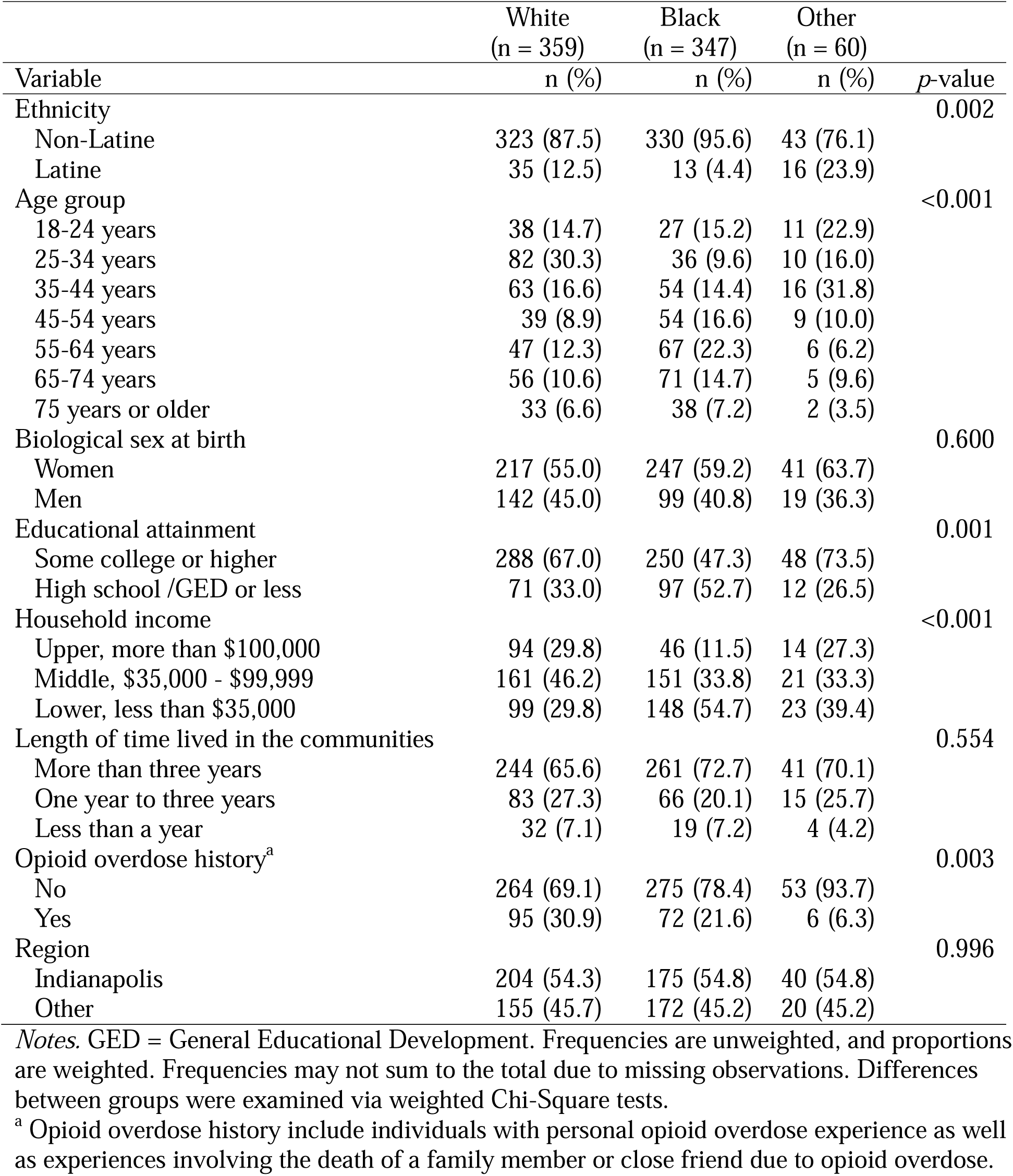
Demographic Characteristics by Race: March-May 2023, (N = 772)

### 3.2. Weighted scores for knowledge on opioid overdose and naloxone administration and perceived competency level to manage opioid overdoses by race

Table 2 presents the unweighted frequencies and weighted proportions of responses to the 10 questions on opioid overdose and naloxone administration knowledge, along with the weighted mean scores of 7 items that assess perceived competency in managing opioid overdose. The average score on the 10 questions assessing knowledge on opioid overdose and naloxone administration showed significant differences by race (*p* < .001). White participants had the highest scores (Mean = 6.65, *SD* = 1.86), followed by other racial groups (Mean = 5.75, *SD* = 1.89), and then Black participants (Mean = 5.70, *SD* = 1.79). In contrast, perceived competency to manage opioid overdose, assessed through 7 survey items, did not show significant variation across racial groups (*p* = 0.478). White participants had a mean score of 2.95 (*SD* = 1.05), Black participants scored 2.82 (*SD* = 0.92), and Other racial groups scored 2.96 (*SD* = 1.07).

**Table 2.** Weighted Scores for Knowledge on Opioid Overdose and Naloxone Administration and Perceived Competency Level to Manage Opioid Overdoses by Race: March-May, 2023 (N = 772)

|  | White<br>(n = 359) | Black<br>(n = 347) | Other<br>(n = 60) |  |
| --- | --- | --- | --- | --- |
|  | Mean (SD) |  |  | p-value<sup>a</sup> |
| *Knowledge on Opioid Overdose and Naloxone Administration (10 Questions),* | 6.65 (1.86) | 5.70 (1.79) | 5.75 (1.89) | <0.001 |
| *Perceived Competency to Manage Opioid Overdose (7 items),* | 2.95 (1.05) | 2.82 (0.92) | 2.96 (1.07) | 0.478 |
| *Weighted Percentage of Correct Response - True or False Statement (6 Questions)* | n (%) |  |  | p-value<sup>b</sup> |
| Naloxone is effective in reversing effects of cocaine overdose. (False) | 157 (43.4) | 96 (30.3) | 24 (43.2) | 0.055 |
| All naloxone products are effective in reversing opioid overdose, including fentanyl-involved opioid overdoses. (True) | 219 (58.6) | 216 (63.8) | 31 (53.2) | 0.446 |
| Naloxone should NOT be used for pregnant women in life-threatening opioid overdose circumstances. (False) | 231 (58.5) | 147 (43.0) | 21 (36.6) | 0.012 |
| Because naloxone's effects don't last long, overdose symptoms may come back. (True) | 230 (68.7) | 192 (56.7) | 36 (54.9) | 0.082 |
| Naloxone can provoke withdrawal symptoms like fever, irritability, rapid heart rate, sweating, nausea, and vomiting. (True) | 278 (78.0) | 250 (74.1) | 44 (85.4) | 0.253 |
| If the person overdosing does not respond within 2 to 3 minutes after administering a dose of naloxone, a second dose of naloxone can be administered. (True) | 255 (70.6) | 216 (57.4) | 38 (64.8) | 0.070 |
| *Weighted Percentage of Correct Response - Multiple Choices Questions (4 Questions)* | n (%) |  |  | p-value<sup>b</sup> |
| Which of the following is NOT a sign or symptom of opioid overdose? | 229 (66.8) | 190 (52.6) | 26 (39.4) | 0.005 |
| a. Slow or shallow breathing |  |  |  |  |
| b. Having blood-shot eyes (Correct) |  |  |  |  |
| c. Unconsciousness |  |  |  |  |
| d. Very small pupils |  |  |  |  |
| e. Fingernails or lips turning blue/purple |  |  |  |  |
| Which step is NOT recommended when someone has overdosed on opioids? | 302 (86.3) | 255 (72.3) | 51 (88.4) | 0.007 |
| a. Make the person vomit the drugs that may have been swallowed (Correct) |  |  |  |  |
| b. Call 911 for help |  |  |  |  |
| c. Monitor what is happening to the person until an ambulance arrives |  |  |  |  |
| d. Be sure the person's airway is clear |  |  |  |  |
| How should you administer naloxone? | 202 (53.2) | 215 (62.5) | 29 (55.6) | 0.282 |
| a. Spray through the nose |  |  |  |  |
| b. Inject directly into the muscle |  |  |  |  |
| c. Inject directly under the skin |  |  |  |  |
| d. All of them are correct routes of administration (Correct) |  |  |  |  |
| Which is a CORRECT statement? | 358 (81.2) | 340 (66.6) | 58 (54.3) | 0.002 |
| a. There is no need to call for an ambulance if I know how to manage an overdose. |  |  |  |  |
| b. After recovering from an opioid overdose, the person must not take any opioid, but it is okay for them to drink alcohol or take sleeping tablets. |  |  |  |  |
| c. You should not give naloxone more than once to people who suffer from a fentanyl-involved overdose and taking breaths that are slower and shallower than normal. |  |  |  |  |
| d. Someone can overdose again even after having received naloxone. (Correct) |  |  |  |  |

| *Weighted Mean of Each Perceived Competency to Manage Opioid Overdose Item* | *Mean (SD)* |  |  | *p-value<sup>a</sup>* |
| --- | --- | --- | --- | --- |
| 1. I would be able to respond effectively with an overdose | 3.02 (1.35) | 2.74 (1.41) | 3.21 (1.50) | 0.110 |
| 2. I am confident that I can administer naloxone to someone who has overdosed | 3.00 (1.47) | 3.18 (1.41) | 3.13 (1.47) | 0.579 |
| 3. If someone overdoses, I would know what to do to help them | 3.20 (1.28) | 3.01 (1.36) | 3.36 (1.47) | 0.282 |
| 4. I know very little about how to help someone who has overdosed | 2.82 (1.38) | 2.64 (1.44) | 2.65 (1.49) | 0.530 |
| 5. I would be afraid of doing something wrong in an overdose situation | 2.38 (1.33) | 2.41 (1.36) | 2.29 (1.34) | 0.915 |
| 6. I would be afraid of giving naloxone in case the person becomes aggressive afterwards | 3.14 (1.35) | 2.83 (1.34) | 3.00 (1.11) | 0.153 |
| 7. If I tried to help someone who has overdosed, I might accidentally hurt them | 3.09 (1.18) | 2.97 (1.29) | 3.11 (1.34) | 0.701 |

*Notes.* Frequencies may not sum to the total due to missing observations.
<sup>a</sup> Corresponding p-values were calculated via weighted Chi-square tests; <sup>b</sup> Corresponding p-values were calculated via weighted t-tests.

Accordingly, some individual knowledge questions (components of the overall knowledge score) showed significant variation by racial group, whereas none of the individual weighted mean scores for perceived competence items (components of the overall weighted mean competency score) indicated significant differences among racial groups.

### 3.3. Factors associated with knowledge on opioid overdose and naloxone administration scores

Table 3 presents the multilevel analysis of factors influencing knowledge about opioid overdose and naloxone administration. The model included both individual and zip code-level predictors, as well as interaction terms. In the model, Black (= −1.74, *p* = 0.005) participants had significantly lower opioid overdose and naloxone administration knowledge scores compared to their White counterparts. Education level also played a significant role, with participants having a high school education or less scoring lower than those with some college or higher education (= −0.31, *p* = 0.044). Income was another significant predictor; compared to those earning > $100,000, those earning < $100,000 showed lower knowledge scores (= −0.47, *p* = 0.009 for those earning $35,000 – $99,999 and = −0.66, *p* = 0.002 for those earning less than $35,000). Notably, participants with a history of opioid overdose demonstrated higher knowledge scores (= 0.83, *p* = <0.001). For the length of time lived in the community, individuals who lived in their area for one to three years scored lower than those who lived there for more than three years (= −0.42, *p* = 0.012). Zip code-level predictors showed no significant relationship with knowledge scores. However, a significant cross-level interaction was found between Black race and FPL<200%, indicating that Black residents living in poorer neighborhoods displayed lower knowledge scores than White counterparts (= 1.06, *p* = 0.039). The intraclass correlation coefficient (ICC) for the null model was 0.028, indicating that 2.8% of the total variance in the knowledge level was due to difference between the zip codes.

**Table 3.**
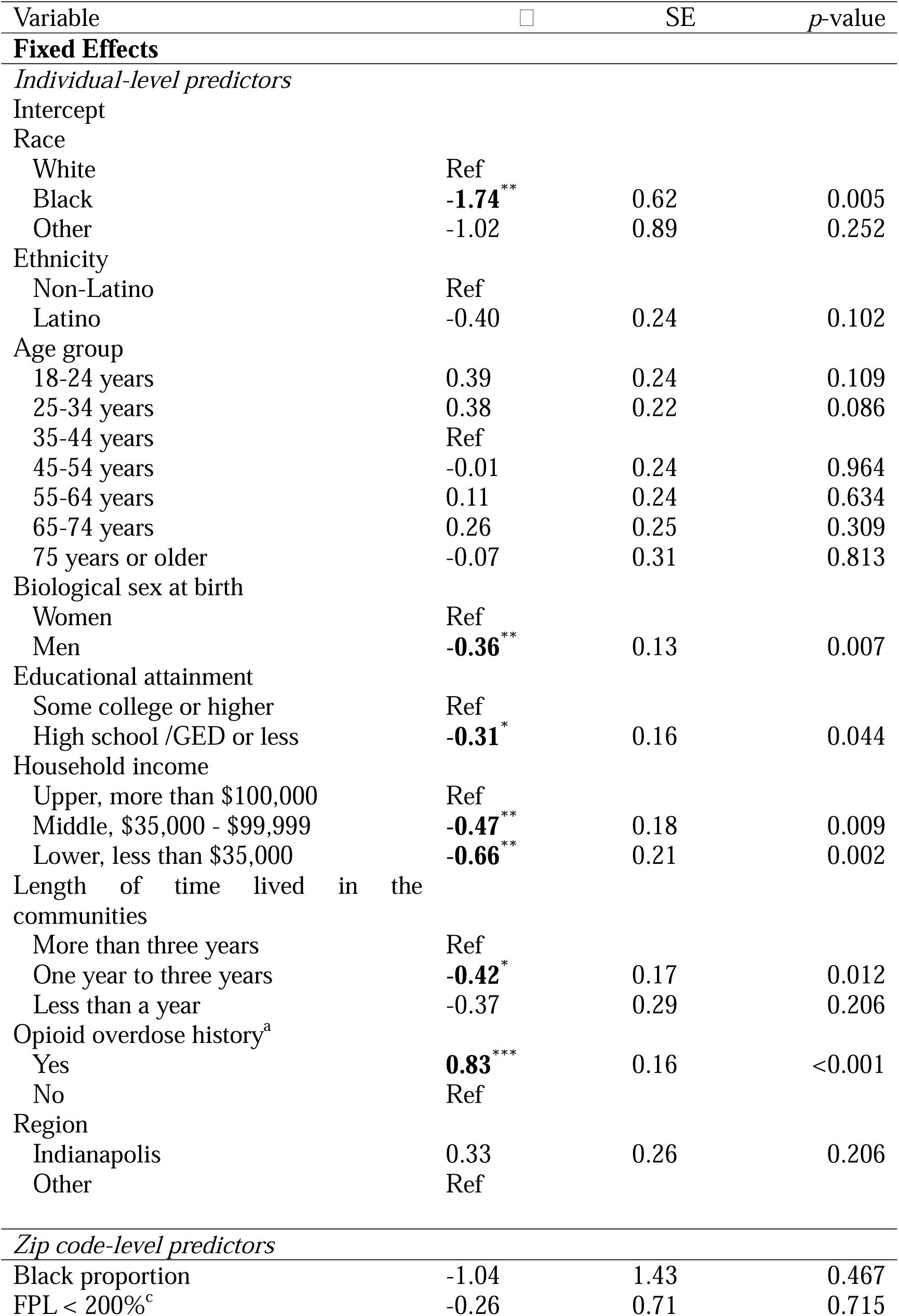
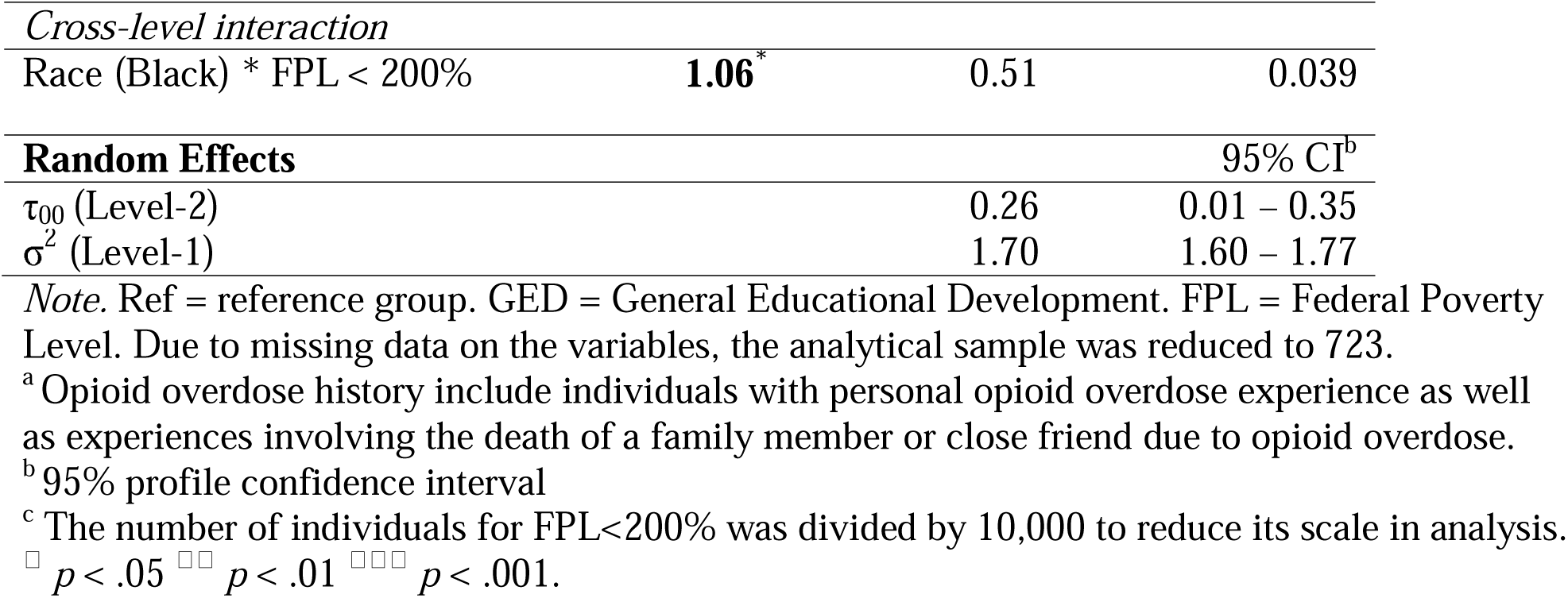
Multilevel Model Analysis of Knowledge on Opioid Overdose and Naloxone.

### 3.4. Factors associated with perceived competency on opioid overdose and naloxone administration

Table 4 summarizes the multilevel model analysis on perceived competency in managing opioid overdose and administering naloxone. At the individual level, race was not a significant predictor, but age was. The youngest age group (18-24 years) reported significantly lower perceived competency (β = −0.34, *p* = 0.013) compared to the 35-44 age group, as did the oldest groups, aged 65-74 (β = −0.45, *p* = 0.001) and 75+ (β = −0.33, *p* = 0.013). No significant differences were noted for the other age groups. The only other significant predictor of perceived competency was having a personal or familial history of opioid overdose, which was associated with a higher score (β = 0.61, *p* < 0.001). The ICC of the null model was 0.004, indicating that only 0.4% of the variance in perceived competency was due to differences between zip codes. Random effects analysis showed minimal variability across zip codes.

**Table 4.** Multilevel Model Analysis of Perceived Competency on Opioid Overdose and Naloxone Administration (N=723)

| Variable | $\beta$ | SE | p-value |
| --- | --- | --- | --- |
| **Fixed Effects** |  |  |  |
| *Individual-level predictors* |  |  |  |
| Intercept |  |  |  |
| Race |  |  |  |
| White | Ref |  |  |
| Black | -0.58 | 0.29 | 0.093 |
| Other<sup>a</sup> | 0.63 | 0.49 | 0.203 |
| Ethnicity |  |  |  |
| Non-Latine | Ref |  |  |
| Latine | -0.25 | 0.13 | 0.068 |
| Age group |  |  |  |
| 18-24 years | **-0.34\*** | 0.13 | 0.013 |
| 25-34 years | -0.11 | 0.12 | 0.392 |
| 35-44 years | Ref |  |  |
| 45-54 years | -0.15 | 0.14 | 0.264 |
| 55-64 years | -0.04 | 0.13 | 0.764 |
| 65-74 years | **-0.45\*\*** | 0.14 | 0.001 |
| 75 years or older | **-0.33\*** | 0.17 | 0.013 |
| Biological sex at birth |  |  |  |
| Women | Ref |  |  |
| Men | 0.05 | 0.07 | 0.480 |
| Educational attainment |  |  |  |
| Some college or higher | Ref |  |  |
| High school /GED or less | -0.06 | 0.09 | 0.456 |
| Household income |  |  |  |
| Upper, more than \$100,000 | Ref | | |
| Middle, \$35,000 - \$99,999 | -0.08 | 0.10 | 0.429 |
| Lower, less than \$35,000 | -0.08 | 0.11 | 0.469 |
| Length of time lived in the communities |  |  |  |
| More than three years | Ref |  |  |
| One year to three years | -0.02 | 0.09 | 0.817 |
| Less than a year | -0.09 | 0.16 | 0.571 |
| Opioid overdose history<sup>b</sup> |  |  |  |
| Yes | **0.61\*\*\*** | 0.09 | <0.001 |
| No | Ref |  |  |
| Region |  |  |  |
| Indianapolis | -0.07 | 0.10 | 0.491 |
| Other | Ref |  |  |
| *Zip code-level predictors* |  |  |  |
| Black proportion | 0.24 | 0.55 | 0.664 |
| FPL < 200%<sup>c</sup> | -0.08 | 0.31 | 0.878 |
| *Cross-level interaction* |  |  |  |
| Race (Black) * FPL < 200% | 0.41 | 0.28 | 0.144 |
| **Random Effects** |  |  | 95% CI<sup>b</sup> |
| $\tau_{00}$ (Level-2) | | 0.00 | 0.00 – 0.00 |
| $\sigma^2$ (Level-1) | | 0.90 | 0.89 – 0.99 |

*Note.* Ref = reference group. GED = General Educational Development. FPL = Federal Poverty Level. Due to missing data on the variables, the analytical sample was reduced to 723.
<sup>a</sup> Opioid overdose history include individuals with personal opioid overdose experience as well as experiences involving the death of a family member or close friend due to opioid overdose.
<sup>b</sup> 95% profile confidence interval
<sup>c</sup> The number of individuals for FPL<200% was divided by 10,000 to reduce its scale in analysis.
□ $p < .05$ □□ $p < .01$ □□□ $p < .001$

## 4. Discussion

This study is one of the first to examine the level of knowledge and perceived competency regarding opioid overdose and naloxone administration among racially diverse urban residents in Indiana. We focused on select urban zip codes within Indianapolis, Gary, Merrillville, South Bend, and Fort Wayne, utilizing recent representative data to capture the current landscape. Our findings revealed significant racial differences in opioid overdose and naloxone administration knowledge scores, with White urban residents in Indiana scoring the highest and Black urban residents scoring the lowest. This discrepancy highlights potential disparities in access to or engagement with educational resources related to opioid overdose and naloxone use. Differences in knowledge by race also interacted with community-level poverty, such that Black residents living in high poverty zip codes displayed lower knowledge scores than Black residents in other areas included in the study. The average score of Indiana’s urban residents (across all races) was approximately 6 out of 10 questions correct. The results indicate that there is considerable room for improvement in overall knowledge levels, as well as a need to investigate whether there are racial differences around availability of educational resources regarding opioid overdose and naloxone usage, and, if so, what factors cause those differences.

Our finding that Black urban residents scored lower on knowledge questions around opioid overdose and naloxone administration compared to White urban residents is consistent with prior research (30, 31). Likewise, the relatively low knowledge score across all urban Indiana residents in the sample aligns with previous research indicating that despite naloxone’s relatively widespread availability, knowledge about naloxone is lacking (21, 32–34). This could likely be improved through peer-led educational initiatives (35–39), which are successful in improving participants’ knowledge about opioid overdose and naloxone administration.

Seo et al. (2023) found that Black residents in Indianapolis felt they lacked sufficient educational opportunities regarding opioid overdose and naloxone usage compared to White urban residents (40). However, they expressed a readiness to learn about and carry naloxone for overdose response, particularly when the education and support came from trusted figures within their community such as Black church leaders (40). Addressing the gap observed in this study might be achievable by implementing culturally tailored education programs (41, 42) involving community leaders who grasp the cultural nuances and can communicate effectively with Black residents about the importance of naloxone administration (43).

Also consistent with previous studies (44, 45), our findings indicated that having a history of opioid overdose, whether personal or through the death of a family member or close friend, was associated with significantly higher knowledge scores. This suggests that firsthand experience or proximity to opioid-related events may serve as a catalyst for increased awareness and understanding (44, 45), potentially due to greater engagement with medical professionals or support services following such events. Interestingly, no significant associations were found between zip code-level predictors and knowledge scores except the poverty level of the community. Research has shown that poverty and socioeconomic status are linked to educational access and health literacy (46) thereby affecting health outcomes (47). Additionally, aligning with previous studies (45, 48) several socioedemographic factors were significantly related with knowledge scores. Men, individuals with lower household income and educational attainment, and those in lower-income brackets exhibited lower knowledge scores. Targeted educational interventions, increased resource accessibility, partnerships with local organizations, tailored messaging, financial incentives, and ongoing evaluation are all theoretical strategies that might help address differences in knowledge scores among men, lower-income, and less-educated groups, though whether any specific strategy would be effective in doing so is beyond the scope of this study. Additionally, we found a cross-level interaction between race and poverty level. This interaction emphasizes the need for targeted interventions that address both race and socioeconomic status such that more Black residents in high-poverty areas are provided enhanced educational opportunities on opioid overdose and naloxone administration.

Regarding perceived competency in managing opioid overdose and naloxone administration, age emerged as a significant predictor, with both the youngest (18-24 years) and oldest age groups (65-74 and 75 years or older) reporting lower perceived competency. This may indicate a lack of practical experience or exposure to training opportunities among these cohorts. Notably, having an opioid overdose history significantly enhanced perceived competency, aligning with our previous findings regarding knowledge scores. This relationship suggests that experiential learning may play an important role in developing confidence in overdose management. In a study based in Nebraska, Cooper-Ohm et al. (2023) found an insignificant relationship between past history and familiarity with naloxone in cities of Omaha and Lincoln, however a significant relationship was found in regions outside these two cities (49). Furthermore, the ICC results from the null models in this study provide insights into the variability of knowledge and perceived competency related to opioid overdose management and naloxone administration across different urban communities. Specifically, the ICC showed that 2.8% of the variance in knowledge was due to geographical differences. Urban communities with limited access to resources could benefit from educational campaigns and support services that address their specific needs (50). By acknowledging the community context, public health practitioners could develop more effective strategies to share important information about opioid overdose prevention and naloxone use (35, 51, 52). On the other hand, the ICC for perceived competency was only 0.4%, which indicates that differences in perceived competency across zip codes are minimal, regardless of the availability of educational resources in these communities.

The findings of this study should be cautiously interpreted within the context of its limitations. By focusing on specific zip codes and recruiting survey respondents from only those zip codes, there may be limited generalizability, potentially applying only to the settings with similar characteristics and environments. Nonetheless, the chosen zip codes were deliberately selected from urban areas with higher proportions of Black residents and elevated rates of overdose events and deaths compared to other areas in Indiana, rendering the findings of our study meaningful. Additionally, since the survey relied on self-reported data, it is susceptible to response bias and recall bias. Furthermore, we could not assess temporal relationships due to the cross-sectional nature of the survey. Regarding survey questions on opioid overdose and naloxone, the inclusion of only 10 questions for knowledge and 7 items for perceived competency may not encompass all aspects of knowledge and perceived competency pertaining to opioid overdose and naloxone administration.

Many Indiana residents continue to die from opioid-involved overdose. While the strategies to reduce this rate are many-pronged, ensuring layperson access to naloxone, knowledge around how to use it and recognize an overdose, and self-efficacy to intervene in case of an overdose is likely important components. Our findings suggest that there is a need for widespread and readily accessible education for urban residents to enhance the understanding of opioid overdose and naloxone administration. In providing those educational opportunities, it may be important to further investigate whether they are accessible to Black urban residents, who may have fewer opportunities to receive such training. As a part of the MACRO-B project, our current efforts involve educating the community on opioid overdose, naloxone administration and its distribution. In the future, we plan to re-evaluate these outcomes to gauge the long-term effectiveness and sustainability of our efforts from this project.

## Data Availability

Data are not available due to personally identifiable information of study participants who were recruited from relatively small geographic areas (8 zip code areas) and sensitive data (e.g., illicit drug use).

## Acknowledgements

This work was supported by the Indiana University Center for Survey Research (CSR). We thank the CSR for assisting in administering community surveys in our target study area which greatly assisted the research although they may not agree with all of the interpretations/conclusions of this paper.

## Funding Statement

This project was supported by the Office of Minority Health (OMH) of the U.S. Department of Health and Human Services (HHS) as part of a financial assistance award (CPIMP221346). The contents are those of the authors and do not necessarily represent the official views of, nor an endorsement, by OMH/OASH/HHS, or the U.S. Government. For more information, please visit https://www.minorityhealth.hhs.gov/.

## Conflict of interest

All authors declare that they have no conflicts of interest.

## Financial disclosure

This Project was supported by the Office of Minority Health (OMH) of the U.S. Department of Health and Human Services (HHS) as part of a financial assistance award (CPIMP221346). The contents are those of the authors and do not necessarily represent the official views of, nor an endorsement, by OMH/OASH/HHS, or the U.S. Government. For more information, please visit https://www.minorityhealth.hhs.gov/.

## Notes

### Competing Interest Statement

The authors have declared no competing interest.

